# Primary care metronidazole prescription in public and private facilities of South Benin: A register-based cross-sectional study

**DOI:** 10.64898/2026.04.07.26350314

**Authors:** Tankpinou Zoumenou Harold, Affolabi Dissou, Kohoun Rodrigue, Sandjakian Léna, Faucher Jean-François

**Affiliations:** Clinical Research Institute of Benin (IRCB), Abomey-Calavi, Benin; Supranational Reference Laboratory for Tuberculosis, Cotonou, Benin; Laboratory of Epidemiology of Chronic and Non-Communicable Diseases, Cotonou, Benin; Epimact: Inserm U1094, IRD UMR270, Univ. Limoges, CHU Limoges, EpiMaCT - Epidemiology of chronic diseases in tropical zone, Institute of Epidemiology and Global Health – Michel Dumas, OmegaHealth, Limoges, France; Department of Infectious Diseases and Tropical Medicine, University Hospital of Limoges, 2 Avenue Martin Luther King, 87000 Limoges, France; Coordination of Abomey-Calavi–So-Ava Health Zone, Ministry of Health, Benin; Groupe Vaccination-Prévention de La Société de Pathologie Infectieuse de Langue Française (SPILF), France; CHU Limoges, Department of Infectious Diseases and Tropical Medicine, Limoges, France; Sorbonne Université, Inserm, Institut Pierre-Louis d’épidémiologie et de santé publique, F-75012 Paris, France

**Keywords:** antimicrobials, primary care, public and private facilities, low-income countries, metronidazole

## Abstract

**Background:** Metronidazole (MTZ) is a first-line antibiotic for several enteric infections. Its use is common in low-income countries, where most primary-care consultations are conducted by nurses. However, increasing resistance among some enteric pathogens is a growing concern. Using WHO guidelines, we conducted a register-based cross-sectional study to assess MTZ prescribing practices and their determinants in public and private primary healthcare facilities in South Benin.

**Methods:** We performed a register-based cross-sectional study covering the year 2020 in 11 primary healthcare facilities (5 public and 6 private) in Abomey-Calavi, South Benin, following WHO recommendations. In total, 200 visits per facility were selected using systematic random sampling. The primary outcome was the prevalence of MTZ prescription. Determinants of MTZ prescription were identified using multivariable logistic regression analysis.

**Results:** In total, 2,200 medical visits were analyzed. The median age of patients was 19 years, and 57% were female. Antimalarials were prescribed in 52% of visits. Antibacterial agents were prescribed in the majority of visits, with MTZ being the second most frequently prescribed antibiotic (18%), after aminopenicillins (27%). In multivariable analysis, digestive symptoms (adjusted odds ratio [aOR], 8.65; 95% confidence interval [CI], 6.49–11.6), genitourinary symptoms (aOR, 6.84; 95% CI, 3.18–15.0), and skin lesions (aOR, 2.39; 95% CI, 1.58–3.60) were independently associated with increased odds of MTZ prescription. In contrast, fever (aOR, 0.66; 95% CI, 0.49–0.87), respiratory symptoms (aOR, 0.44; 95% CI, 0.26–0.71), and malaria (aOR, 0.21; 95% CI, 0.15–0.28) were associated with decreased odds. Visits in the private sector were also associated with higher odds of MTZ prescription compared with the public sector (aOR, 2.31; 95% CI, 1.78–3.02).

**Conclusion:** MTZ is the second most commonly prescribed antibiotic in primary care in the study area, with its use largely driven by digestive symptoms. Further studies are needed to assess the appropriateness of this prescription. Additionally, research is warranted to understand better the determinants of higher antimicrobial prescribing in the private healthcare sector.

**Highlights:**

- MTZ is the second most prescribed antibiotic in the study area.
- MTZ prescription is primarily driven by digestive symptoms.
- The private healthcare sector is independently associated with higher MTZ prescription rates.
- Antimicrobial prescribing is generally higher in private healthcare facilities than in public facilities.

## Introduction

Combating antimicrobial resistance (AMR) is a major challenge of the 21^st^ century. It has been estimated that 1.14 million deaths were directly attributable to AMR in 2021, of which 209,000 occurred in Sub-Saharan Africa (1). Beyond mortality and disability, AMR imposes substantial economic burdens, partly due to prolonged hospital stays (2). In primary care settings, AMR is expected to result in an increasing number of infections that are either untreatable or costlier and more difficult to manage (3).

Metronidazole (MTZ) is an antibiotic classified within the World Health Organization (WHO) access group under the Access, Watch, Reserve (AWaRe) antibiotic categorization, which includes first- or second-line agents that provide optimal therapeutic value while minimizing the risk of resistance (4). According to the WHO Model List of Essential Medicines, MTZ is indicated as a first-line treatment for intestinal infections caused by *Clostridioides difficile*, trichomoniasis, intra-abdominal infections (such as peritonitis), and necrotizing fasciitis (5).

As MTZ is a first-line therapy for amoebiasis and giardiasis, its use is likely to be more frequent in settings where infections associated with poor water, sanitation, and hygiene are prevalent, such as in low-income countries. Despite this, relatively limited and heterogeneous data are available regarding MTZ use in low- and middle-income countries (6–15).

Resistance to MTZ has already emerged as a concern, particularly in the treatment of diarrheal diseases caused by *Giardia* species and *Clostridioides difficile*, and may further increase due to irrational prescribing practices (16,17). Although MTZ is not included in the WHO watch group, its rational use warrants close attention. While AMR can arise naturally, it is largely driven by inappropriate antibiotic use (18). Therefore, surveillance of antibiotic consumption and the implementation of antibiotic stewardship programs, including in community settings, are essential components in addressing AMR (18).

Antibiotics are widely prescribed in primary care across low- and middle-income countries, where non-medical prescribers play a significant role in delivering healthcare, particularly in Africa (19,20). In this context, and given the lack of studies specifically examining MTZ use in primary healthcare centers, we conducted a register-based cross-sectional study aimed at evaluating the frequency and determinants of MTZ prescribing in public and private primary healthcare centers in South Benin.

## Materials and Methods

### Study type and setting

This register-based cross-sectional study was conducted from April to May, 2021, in public and private primary healthcare facilities in Abomey-Calavi, a city in southern Benin. All included facilities operate under the supervision of the regional public health administration (formal health sector). The study focused on facilities where prescriptions are predominantly issued by nurses. These centers have limited laboratory capacity, with malaria rapid diagnostic tests (RDTs) being the most widely available diagnostic tool. The selection of facilities is shown in the flowchart in Figure 1.

**Figure 1:**
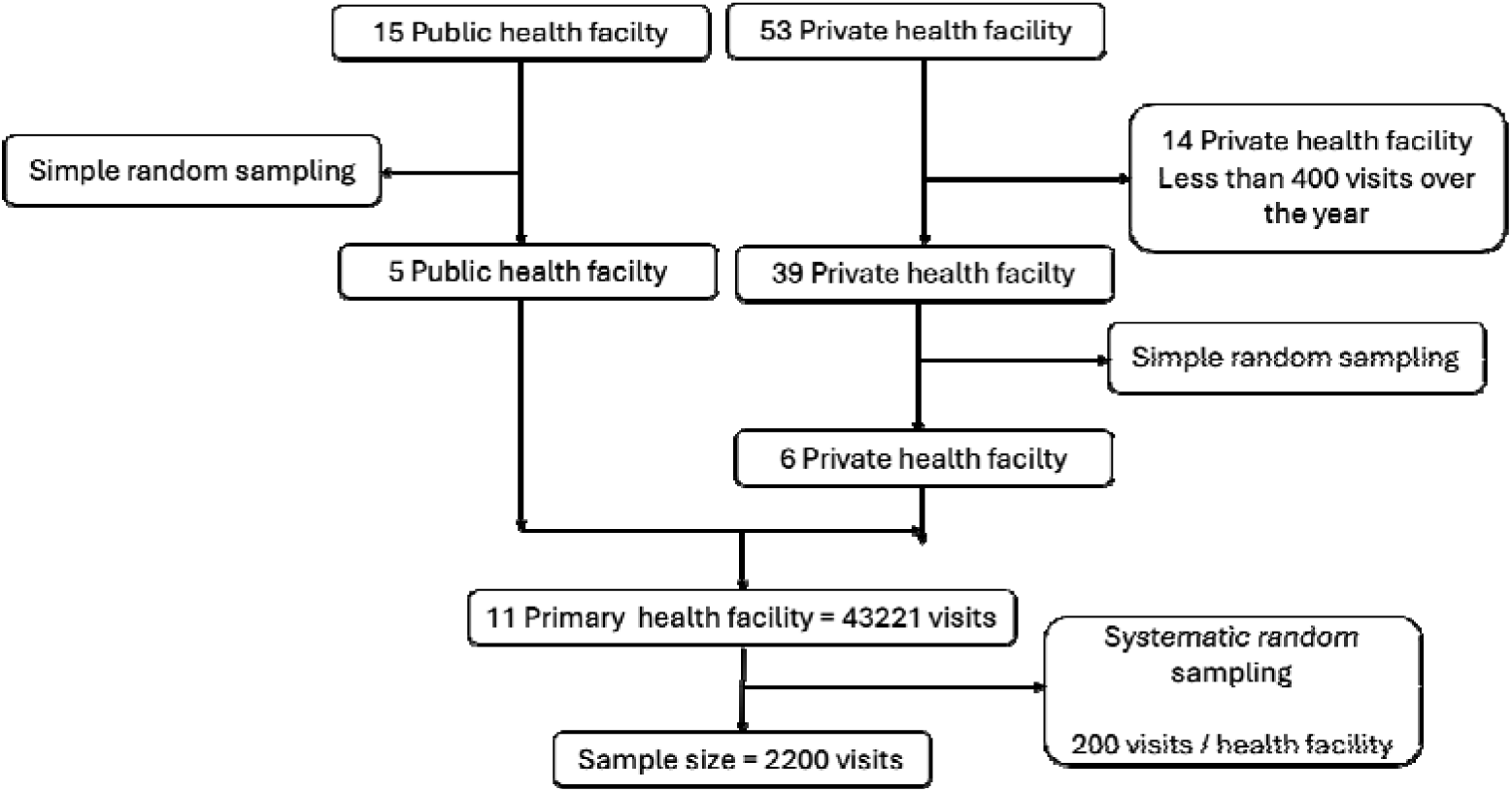
Flowchart of visit selection

### Sample size and inclusion criteria

The sample size was determined in accordance with the WHO guidelines for studies assessing prescribing practices in health facilities (21). The WHO recommends collecting data from at least 600 medical visits across all facilities, with a minimum of 30 visits per selected health center. Thus, the total number of visits depends on the number of selected facilities.

We elected to record 200 medical visits per healthcare facility. Eligible facilities were defined as those belonging to the formal health sector and reporting more than 400 visits per year. All public healthcare facilities met the inclusion criteria, whereas only 39 of the 53 private facilities were eligible. From these, five public healthcare centers and six private healthcare facilities were randomly selected. Based on 200 visits per facility, the final expected sample size was 2,200 medical visits.

### Sampling procedure

A multistage sampling procedure was applied. In the first stage, public and private healthcare facilities were selected as described above.

In the second stage, 200 medical visits per facility were extracted from registries covering the entire year 2020. Systematic random sampling was performed using the registries of recorded visits. The sampling interval was calculated by dividing the total number of annual visits by the number of visits to be collected per facility. The total number of visits per facility was obtained from annual reports provided by the health administration. The first patient was randomly selected between the first entry in the registry and the sampling interval.

### Data collection

Data collected included demographic variables (age, sex, address, etc.), clinical information (symptoms and diagnosis), laboratory investigations (malaria RDT, blood smear, and hemogram), and therapeutic data. Six healthcare providers were trained in data collection using KoboCollect version 2.021.03. Medical visits with missing data were replaced by the next eligible visit while maintaining the predefined sampling interval. The medications analyzed were antibiotics and antimalarials, classified according to the WHO Anatomical Therapeutic Chemical classification system.

### Outcomes

The primary outcome was the prevalence of MTZ prescription.

### Independent variables

Age was recorded as a continuous variable in years, and sex as a binary categorical variable (male or female). For pediatric patients, age in months was converted to years by dividing by 12.

Clinical symptoms were categorized by system and recoded as binary variables. The following categories were defined: digestive symptoms (vomiting, nausea, abdominal pain, oral thrush, or diarrhea), genitourinary symptoms (dysuria, vaginal discharge, urinary burning, urethral discharge, or metrorrhagia), skin lesions (rashes, traumatic or non-traumatic wounds, abscesses, or itching), respiratory symptoms (cough, rhinorrhea, sore throat, or dyspnea), and fever (self-reported and/or documented).

Malaria was considered present if a positive RDT or blood smear was documented, or in the absence of laboratory confirmation, the prescriber recorded malaria as the clinical diagnosis.

### Statistical analysis

Qualitative variables were summarized as counts and percentages, and quantitative variables as medians with interquartile ranges (25th and 75th percentiles). To identify factors associated with metronidazole (MTZ) prescription, we used a two-step modelling strategy. Candidate predictors included demographic characteristics and clinical presentation variables deemed relevant to prescribing decisions. First, a logistic regression model with a least absolute shrinkage and selection operator (LASSO) penalty was applied to select relevant predictors. The optimal penalty parameter (λ) was determined using 10-fold cross-validation, and the value corresponding to the most regularized model within one standard error of the minimum cross-validated error (λ1-SE) was selected. Second, all variables with a non-zero coefficient in the final LASSO model were included in a standard multivariable logistic regression model to estimate adjusted odds ratios and 95% confidence intervals. As a sensitivity analysis, a mixed-effects logistic regression model including a random intercept for study site was fitted to account for potential clustering by site.

### Administrative and ethical aspects

Ethical clearance to access registry data was obtained from the health zone office (No. 58/2021/NS/DDS ATL/ZS AS/BZ/CAR/RRH/SA). The physician or nurse in charge of each primary care facility provided approval for participation. All data were anonymized, and access to the registries was restricted to study personnel.

## Results

### Characteristics of the study population and antimicrobial prescription

Among the 43,221 medical visits recorded in 2020, in total, 2,200 visits from 11 healthcare centers were analyzed, representing 5% of all visits (Figure 1).

As shown in eTable 1 (Supplementary Material), most visits occurred in the private sector. The median age of the study population was 19 years, and 57% of patients were female. Fever was the most common symptom (67%), followed by digestive symptoms (38%) and respiratory symptoms (12%). Antimalarials were prescribed in 52% of visits. Antibacterial agents were also frequently prescribed, including penicillins (27%), MTZ (18%), ciprofloxacin (12%), and ceftriaxone (4.7%).

As shown in eTable 2 (Supplementary Material), malaria testing was frequently documented among visits with fever. An RDT was recorded in 1,044 of 1,453 visits (72%) and was positive in 798 of 1,008 visits with available results (79%). A thick blood smear was documented in 278 of 1,453 visits (19%) and was positive in 98 of 142 visits with available results (69%). Overall, 887 of 1,137 visits with fever and at least one available malaria test result (78%) were positive.

As shown in eTable 3, antimalarial monotherapy was the most common treatment pattern, accounting for 25% of visits. Overall, 20.1% of visits did not receive any antimicrobial therapy. Combination therapies were also frequent: 27.5% of visits received an antimalarial plus at least one antibiotic, while 27.4% received at least one antibiotic without antimalarial therapy.

### Characteristics of the study population and MTZ prescription

As shown in Table 1, visits with MTZ prescription were more frequently associated with digestive symptoms, skin lesions, and genitourinary symptoms compared with visits without MTZ prescription. In contrast, fever (including malaria) and respiratory symptoms were less frequent in visits with MTZ prescription. Demographic characteristics did not differ significantly between the two groups. MTZ prescription was more frequent in the private healthcare sector than in the public sector.

**Table 1:** Population characteristics according to MTZ prescription in public and private healthcare facilities in South Benin, 2021.

| **Variables** | **Metronidazole** |  |  |
| --- | --- | --- | --- |
|  | **No = 1801<sup>1</sup>** | **Yes = 399<sup>1</sup>** | **p-value<sup>2</sup>** |
| Age | 19 (6, 32) | 22 (7, 33) | 0.2 |
| Sex, Female | 1,041 / 1,801 (58%) | 213 / 399 (53%) | 0.11 |
| Public sector | 851 / 1,801 (47%) | 149 / 399 (37%) | <0.001 |

| Variables | Metronidazole |  |  |
| --- | --- | --- | --- |
|  | No = 1801<sup>1</sup> | Yes = 399<sup>1</sup> | p-value<sup>2</sup> |
| Fever | 1,263 / 1,801 (70%) | 190 / 399 (48%) | <0.001 |
| Digestive symptoms | 545 / 1,801 (30%) | 255 / 399 (64%) | <0.001 |
| Respiratory tract symptoms | 239 / 1,801 (13%) | 21 / 399 (5.3%) | <0.001 |
| Skin lesions | 117 / 1,801 (6.5%) | 50 / 399 (13%) | <0.001 |
| Genitourinary symptoms | 14 / 1,801 (0.8%) | 20 / 399 (5.0%) | <0.001 |
| Malaria | 1,088 / 1,801 (60%) | 126 / 399 (32%) | <0.001 |

<sup>1</sup>Median (Q1, Q3); n / N (%)
<sup>2</sup>Wilcoxon rank sum test; Pearson's Chi-squared test

The antimicrobials most frequently co-prescribed with MTZ were antimalarials, ciprofloxacin, and amoxicillin (Table 2). Antimalarial prescription was more common among visits without MTZ prescription than among those with MTZ prescription. Compared to visits without MTZ prescription, those with MTZ prescription had higher rates of co-prescription with ciprofloxacin (32% vs. 7.0%), cotrimoxazole (14% vs. 2.9%), and cloxacillin (8.0% vs. 2.4%) (Table 2).

**Table 2:**
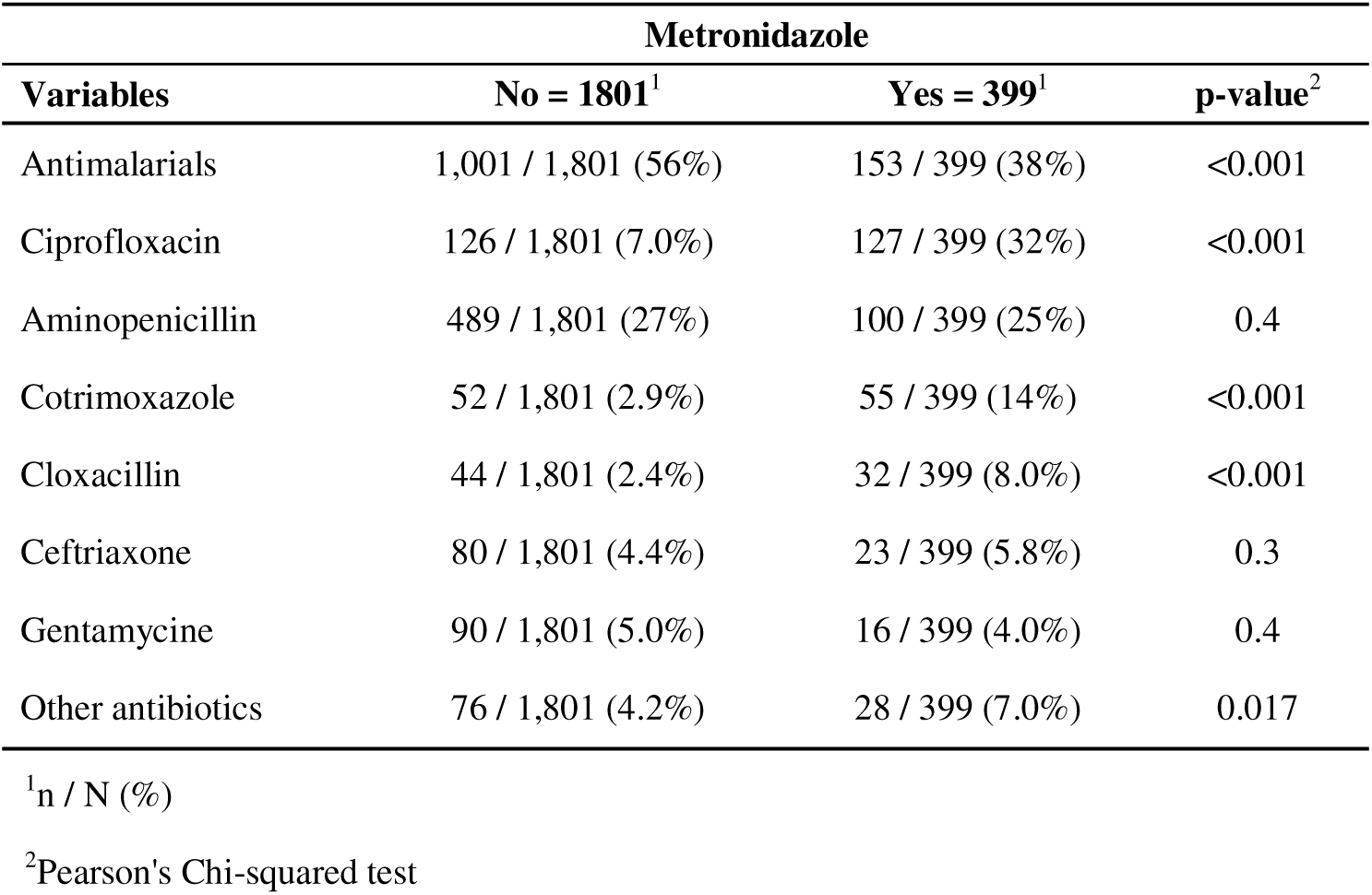
Antimicrobial prescription according to MTZ prescription in public and private healthcare facilities in South Benin, 2021.

### Factors associated with MTZ prescription

As shown in Table 3, digestive symptoms (adjusted odds ratio [aOR], 8.65; 95% confidence interval [CI], 6.49–11.6), skin lesions (aOR, 2.39; 95% CI, 1.58–3.60), and genitourinary symptoms (aOR, 6.84; 95% CI, 3.18–15.0) were independently associated with MTZ prescription in multivariable analysis. Conversely, fever (aOR, 0.66; 95% CI, 0.49–0.87), malaria (aOR, 0.21; 95% CI, 0.15–0.28), and respiratory symptoms (aOR, 0.44; 95% CI, 0.26–0.71) were inversely associated with MTZ prescription.

**Table 3:** Factors independently associated with MTZ prescription in public and private healthcare facilities in South Benin, 2021.

| Variables | Univariate |  | Multivariate |  |
| --- | --- | --- | --- | --- |
|  | cOR(95% CI) | p-value | aOR(95% CI) | p-value |
| Age | 1.00 (1.00, 1.01) | 0.3 |  |  |
| Sex, Female |  |  |  |  |
| Yes | — |  |  |  |
| No | 1.20 (0.96, 1.49) | 0.11 |  |  |
| Public sector |  |  |  |  |
| Yes | — |  | — |  |
| No | 1.50 (1.20, 1.88) | <0.001 | 2.31 (1.78, 3.02) | <0.001 |
| Fever |  |  |  |  |
| No | — |  | — |  |
| Yes | 0.39 (0.31, 0.48) | <0.001 | 0.66 (0.49, 0.87) | 0.003 |
| Digestive symptoms |  |  |  |  |
| No | — |  | — |  |
| Yes | 4.08 (3.26, 5.13) | <0.001 | 8.65 (6.49, 11.6) | <0.001 |
|  | cOR(95% CI) | p-value | aOR(95% CI) | p-value |
| Respiratory tract symptoms |  |  |  |  |
| No | — |  | — |  |
| Yes | 0.36 (0.22, 0.56) | <0.001 | 0.44 (0.26, 0.71) | 0.001 |
| Skin lesions |  |  |  |  |
| No | — |  | — |  |
| Yes | 2.06 (1.44, 2.91) | <0.001 | 2.39 (1.58, 3.60) | <0.001 |
| Genitourinary symptoms |  |  |  |  |
| No | — |  | — |  |
| Yes | 6.74 (3.40, 13.7) | <0.001 | 6.84 (3.18, 15.0) | <0.001 |
| Malaria |  |  |  |  |
| No | — |  | — |  |
| Yes | 0.30 (0.24, 0.38) | <0.001 | 0.21 (0.15, 0.28) | <0.001 |

Abbreviations: CI = Confidence Interval, OR = Odds Ratio

Demographic characteristics were not associated with MTZ prescription. In contrast, healthcare sector was independently associated with MTZ prescription, with higher odds observed in the private sector than in the public sector (aOR, 2.31; 95% CI, 1.78–3.02).

### Sensitivity analyses

eTable 4 (Supplementary) presents the results of the random-effects model used to account for site-level effects. This model did not alter the conclusions of the primary analysis.

### Factors associated with MTZ prescription within sectors: subgroup analysis

To assess whether the determinants of MTZ prescription differed by sector, a subgroup analysis was conducted. As shown in eTable 5, patient characteristics were broadly similar between sectors: age distribution was comparable, females were slightly more represented in the public sector, and malaria was more frequent in the private sector. Digestive symptoms, skin lesions, and genitourinary symptoms were similarly distributed across sectors.

In sector-specific multivariable analyses (eTable 6), malaria was inversely associated with MTZ prescription in both sectors, with a stronger association in the public sector than in the private sector (aOR, 0.11; 95% CI, 0.06–0.18 vs. 0.27; 95% CI, 0.18–0.39). Digestive symptoms, skin lesions, and genitourinary symptoms were positively associated with MTZ prescription in both sectors, with stronger associations observed in the public sector.

## Discussion

In this study setting, MTZ was the second most frequently prescribed antibiotic, and its use was primarily associated with digestive symptoms. In addition to clinical presentation, the private healthcare sector emerged as an independent factor associated with MTZ prescription. Antimalarials and antibacterial agents were widely prescribed across visits, many of which were related to fever. Co-prescription, both between antimalarials and antibiotics and among antibiotics, was common.

This study, based on a rigorous sampling procedure, was specifically designed to provide a reliable description of antimicrobial prescribing practices by nurses in primary care outpatient settings, including both private and public facilities. The retrospective design minimized potential interference between the study objectives and prescribing behavior.

However, several limitations should be acknowledged. First, due to the retrospective design, clinical data were largely limited to basic records of patients’ complaints rather than standardized diagnoses, which were generally unavailable or unreliable except for malaria. Second, antibiotic prescription does not necessarily reflect actual antibiotic consumption, as data on drug dispensing and patient adherence were not available. Finally, data collection occurred during the early phase of the corona virus disease 2019 (COVID-19) pandemic, which may have influenced consultation rates and prescribing practices. However, chloroquine, which was incorporated into COVID-19 management in 2020, was restricted to dedicated diagnostic centers and prescribed only for confirmed cases, limiting its impact on the present findings.

Providing reliable data on antibiotic prescribing in primary care is essential. Our results indicate that MTZ prescription is common in the study area, accounting for 18% of all medical visits. In contrast, in primary care settings in France, MTZ represents less than 1.9% of antibiotic prescriptions, with similar trends observed in England (22,23). Thus, the proportion of MTZ prescriptions in this study was significantly higher than in high-income settings.

Data from low- and middle-income countries remain heterogeneous. In hospital-based studies of inpatient antibiotic use, MTZ was not reported in a study conducted in South India (6), whereas it was among the most frequently prescribed antibiotics in studies from Africa (7–9) and Nepal (10). In a tertiary hospital in Nigeria, nitroimidazoles were the second most commonly prescribed antibiotics after penicillins among outpatients (11). In community settings, MTZ was not reported in a survey from South India (12) and ranked only seventh in Timor-Leste (13), but was commonly used to treat acute gastroenteritis in Nigeria (14) and frequently used for self-medication for respiratory symptoms in Nepal (15).

This study specifically examined the determinants of MTZ prescription. Digestive symptoms, one of the most common reasons for consultation, were the primary driver of MTZ use. This high frequency of MTZ prescription likely reflects a tendency among nurses to suspect enteric infections, which are often treated with MTZ in this context. The frequent co-prescription of ciprofloxacin, a commonly used antibiotic for enteric bacterial infections, further supports this interpretation. However, data on the epidemiology of enteric pathogens and the effectiveness of MTZ against these pathogens in the study setting are limited. Therefore, antibiotic stewardship studies focusing on MTZ use are needed to determine the proportion of enteric infections caused by pathogens susceptible to MTZ and the clinical effectiveness of MTZ in this context.

The observed association between MTZ prescription and skin lesions was unexpected. MTZ is recommended only for severe soft tissue infections and should be administered prior to referral according to local treatment guidelines (ordinogramme) (Additional Files). However, most recorded skin lesions were traumatic wounds, and available data did not allow confirmation of infection. Prospective studies assessing the knowledge, attitudes, and practices of nurses regarding the management of skin infections would be valuable for optimizing antibiotic use.

In contrast, the inverse association between MTZ prescription and malaria or fever was consistent with previous findings showing that the use of RDT-based malaria diagnosis in the management of febrile illness is associated with reduced antibiotic prescribing (24).

The only characteristic independently associated with MTZ prescription that was unrelated to clinical status was the healthcare sector. This finding prompted additional analyses focusing specifically on sector-related differences. Overall, antibiotic prescription (eTable 2), including of MTZ, was more frequent in the private sector. Patient characteristics differed only marginally between private and public sectors with respect to MTZ prescription. In both sectors, digestive symptoms, skin lesions, and genitourinary symptoms were associated with increased MTZ use. Malaria was associated with lower odds of MTZ prescription in both sectors; however, this association was weaker in the private sector. This implies that the propensity to prescribe MTZ remains relatively high in private facilities, even during visits associated with malaria.

The higher rate of antibiotic prescribing in the private sector may be explained by several factors. First, nurses in the public sector are typically supervised by medical doctors, whereas those in the private sector may practice with less oversight. In addition, drug dispensing in the public sector is generally regulated by a medical officer, which is not always the case in private facilities. Patients seeking care in the private sector may also be perceived as having greater financial resources, potentially encouraging increased drug prescribing. Finally, financial incentives associated with prescribing practices may differ between sectors (25,26). Further research is needed to determine whether the higher rate of anti-infective prescribing in the private sector represents a broader trend, particularly in West Africa. If confirmed, studies aimed at identifying the determinants of antibiotic overprescription in private healthcare settings would be of considerable importance.

## Conclusions

This study demonstrated that MTZ is commonly prescribed in primary care in the study area, with its use primarily driven by the presence of digestive symptoms. Further research is warranted to assess the appropriateness of MTZ indications among patients presenting with digestive complaints in this setting. An additional and unexpected finding was the higher rate of prescribing in the private healthcare sector, highlighting the need for targeted investigations to elucidate its underlying determinants.

## Data Availability

All data produced in the present study are available upon reasonable request to the authors

## Authors’ Contributions

Harold Tankpinou and Jean-François Faucher conceived and designed the study, interpreted the results, and drafted the manuscript. Harold Tankpinou performed the data analysis. Dissou Affolabi and Rodrigue Kohoun contributed to field implementation and data collection and critically revised the manuscript. Léna Sandjakian drafted the manuscript. All authors read and approved the final version of the manuscript.

## Acknowledgments

The authors would like to express their gratitude to the Région Nouvelle-Aquitaine for financial support and to the NeuroCM project for additional funding. They also thank the EpiMaCT Laboratory (Epidemiology of Chronic Diseases in Tropical Zones, University of Limoges) for its scientific and logistical support.

The authors are grateful to Josselin Brisset and Patrick Imbert for their review and constructive comments, which helped improve the quality of this study.

Special thanks are extended to the Abomey-Calavi–So-Ava Health Zone and to all public and private primary healthcare centers that agreed to participate in the study. The authors also sincerely thank the healthcare professionals and survey staff who contributed to the successful completion of this study.

## Availability of data and materials

The datasets used and analyzed during the current study are available from the corresponding author on reasonable request.

## Appendix

**eTable 1:** Description of the study population and antimicrobial prescription in public and private healthcare facilities in South Benin, 2021.

| Variables | N = 2,200<sup>1</sup> |
| --- | --- |
| Age | 19 (6, 32) |
| Sex, Female | 1,254 / 2,200 (57%) |
| Public sector | 1,000 / 2,200 (45%) |
| Fever | 1,453 / 2,200 (66%) |
| Digestive symptoms | 800 / 2,200 (36%) |
| Respiratory tract symptoms | 260 / 2,200 (12%) |
| Skin lesions | 167 / 2,200 (7.6%) |
| Genitourinary symptoms | 34 / 2,200 (1.5%) |
| Malaria | 1,214 / 2,200 (55%) |
| Antimalarials | 1,154 / 2,200 (52%) |
| Aminopenicillin | 589 / 2,200 (27%) |
| Metronidazole | 399 / 2,200 (18%) |
| Ciprofloxacin | 253 / 2,200 (12%) |
| Cotrimoxazole | 107 / 2,200 (4.9%) |
| Gentamycine | 106 / 2,200 (4.8%) |
| Ceftriaxone | 103 / 2,200 (4.7%) |
| Cloxacillin | 76 / 2,200 (3.5%) |
| Other antibiotics | 104 / 2,200 (4.7%) |

<sup>1</sup>Median (Q1, Q3); n / N (%)

**eTable 2:** Fever, malaria, and malaria diagnostic testing in public and private healthcare facilities in South Benin, 2021.

| Variables | Fever |  |  | p-value<sup>2</sup> |
| --- | --- | --- | --- | --- |
|  | No = 747<sup>1</sup> | Yes = 1453<sup>1</sup> | Overall<sup>1</sup> |  |
| Malaria | 181 / 747 (24%) | 1,033 / 1,453 (71%) | 1,214 / 2,200 (55%) | <0.001 |
| Rapid diagnostic test (RDT) documented | 218 / 747 (29%) | 1,044 / 1,453 (72%) | 1,262 / 2,200 (57%) | <0.001 |
| Rapid diagnostic test (RDT) positive<sup>3</sup> | 105 / 190 (55%) | 798 / 1,008 (79%) | 903 / 1,198 (75%) | <0.001 |
| Thick blood smear documented | 102 / 747 (14%) | 278 / 1,453 (19%) | 380 / 2,200 (17%) | 0.001 |
| Thick blood smear positive<sup>3</sup> | 27 / 39 (69%) | 98 / 142 (69%) | 125 / 181 (69%) | >0.9 |
| Any test positive (RDT or Smear) | 129 / 223 (58%) | 887 / 1,137 (78%) | 1,016 / 1,360 (75%) | <0.001 |

<sup>1</sup>n / N (%)
<sup>2</sup>Pearson's Chi-squared test
<sup>3</sup>Percentages for malaria diagnostic tests were calculated among patients with an available result for the respective test. Patients without an available result were excluded from the denominator.

**eTable 3:** Patterns of antimicrobial combination therapy in public and private healthcare facilities in South Benin, 2021.

| Variables | N = 2,200<sup>1</sup> |
| --- | --- |
| Pattern of antimicrobial combination |  |
| No antimicrobial therapy | 443 (20.1%) |
| Antimalarial only | 549 (25.0%) |
| Antimalarial + 1 antibiotic | 389 (17.7%) |
| Antimalarial + ≥2 antibiotics | 216 (9.8%) |
| 1 antibiotic without antimalarial | 364 (16.5%) |
| ≥2 antibiotics without antimalarial | 239 (10.9%) |

<sup>1</sup>n (%)

**eTable 4:** Factors independently associated with MTZ prescription in a mixed-effects logistic regression model accounting for center-level clustering.

| Variables | aOR (95% CI) | p-value |
| --- | --- | --- |
| Public sector |  |  |
| Yes | — |  |
| No | 2.39 (1.48, 3.86) | <0.001 |
| Fever |  |  |
| No | — |  |
| Yes | 0.70 (0.53, 0.94) | 0.018 |
| Digestive symptoms |  |  |
| No | — |  |
| Yes | 9.10 (6.75, 12.3) | <0.001 |
| Respiratory tract symptoms |  |  |
| No | — |  |
| Yes | 0.43 (0.26, 0.71) | 0.001 |
| Skin lesions |  |  |
| No | — |  |
| Yes | 2.36 (1.55, 3.59) | <0.001 |
| Genitourinary symptoms |  |  |
| No | — |  |
| Yes | 6.89 (3.12, 15.2) | <0.001 |
| Malaria |  |  |
| No | — |  |
| Yes | 0.19 (0.14, 0.26) | <0.001 |

Abbreviations: CI = Confidence Interval, OR = Odds Ratio

**eTable 5:**
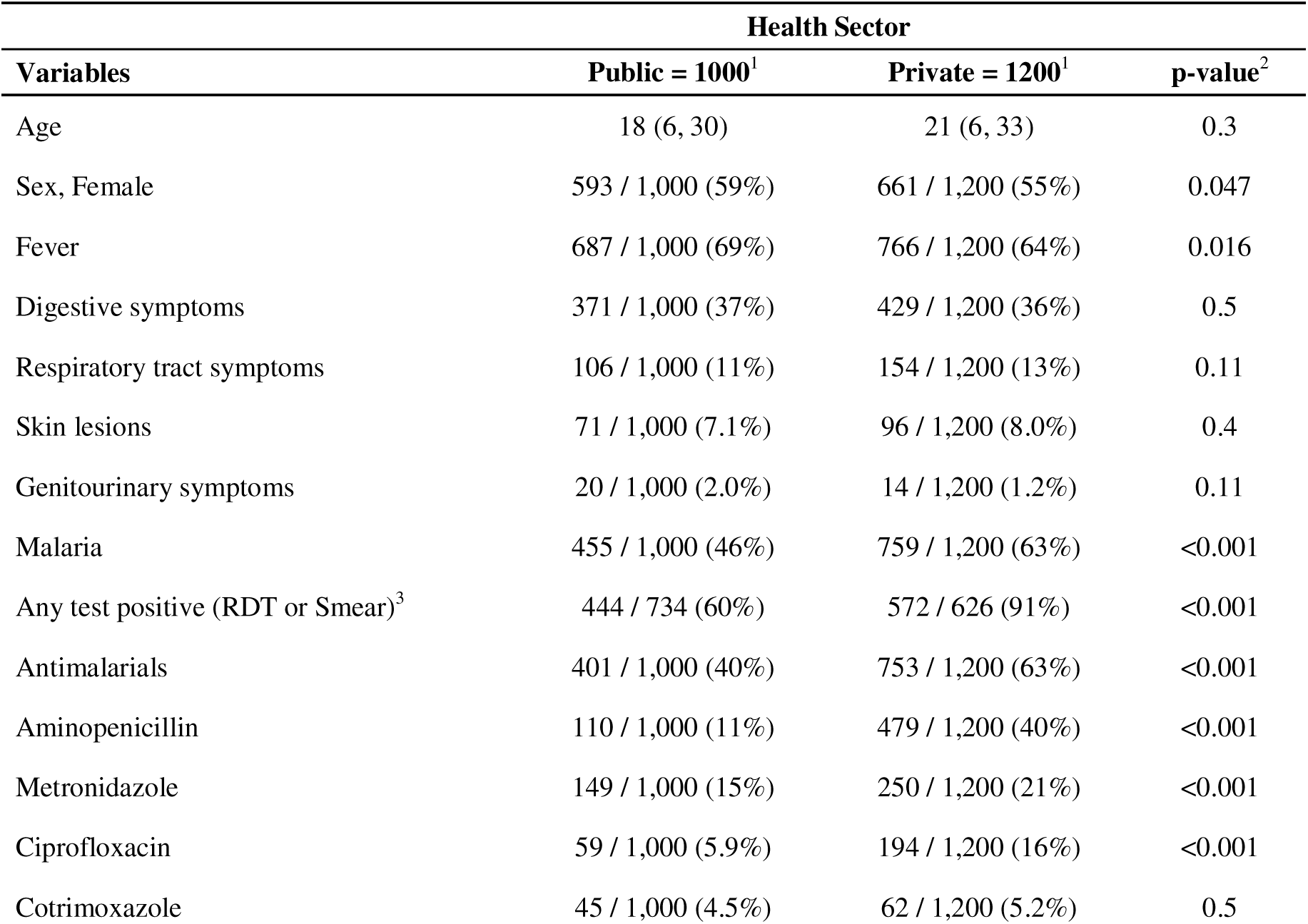
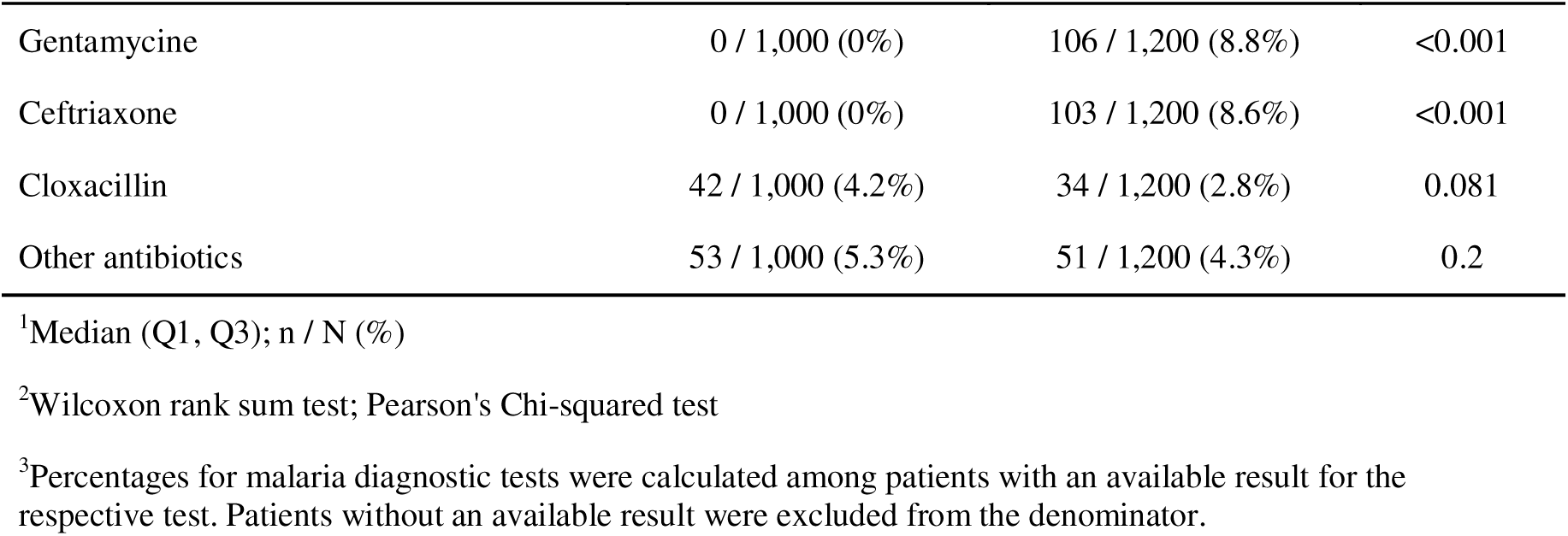
Description of the study population according to healthcare sector in public and private healthcare facilities in South Benin, 2021.

| Variables | Health Sector |  |  |
| --- | --- | --- | --- |
|  | Public = 1000<sup>1</sup> | Private = 1200<sup>1</sup> | p-value<sup>2</sup> |
| Age | 18 (6, 30) | 21 (6, 33) | 0.3 |
| Sex, Female | 593 / 1,000 (59%) | 661 / 1,200 (55%) | 0.047 |
| Fever | 687 / 1,000 (69%) | 766 / 1,200 (64%) | 0.016 |
| Digestive symptoms | 371 / 1,000 (37%) | 429 / 1,200 (36%) | 0.5 |
| Respiratory tract symptoms | 106 / 1,000 (11%) | 154 / 1,200 (13%) | 0.11 |
| Skin lesions | 71 / 1,000 (7.1%) | 96 / 1,200 (8.0%) | 0.4 |
| Genitourinary symptoms | 20 / 1,000 (2.0%) | 14 / 1,200 (1.2%) | 0.11 |
| Malaria | 455 / 1,000 (46%) | 759 / 1,200 (63%) | <0.001 |
| Any test positive (RDT or Smear)<sup>3</sup> | 444 / 734 (60%) | 572 / 626 (91%) | <0.001 |
| Antimalarials | 401 / 1,000 (40%) | 753 / 1,200 (63%) | <0.001 |
| Aminopenicillin | 110 / 1,000 (11%) | 479 / 1,200 (40%) | <0.001 |
| Metronidazole | 149 / 1,000 (15%) | 250 / 1,200 (21%) | <0.001 |
| Ciprofloxacin | 59 / 1,000 (5.9%) | 194 / 1,200 (16%) | <0.001 |
| Cotrimoxazole | 45 / 1,000 (4.5%) | 62 / 1,200 (5.2%) | 0.5 |
|  | Public = 1000<sup>1</sup> | Private = 1200<sup>1</sup> | p-value<sup>2</sup> |
| Gentamycine | 0 / 1,000 (0%) | 106 / 1,200 (8.8%) | <0.001 |
| Ceftriaxone | 0 / 1,000 (0%) | 103 / 1,200 (8.6%) | <0.001 |
| Cloxacillin | 42 / 1,000 (4.2%) | 34 / 1,200 (2.8%) | 0.081 |
| Other antibiotics | 53 / 1,000 (5.3%) | 51 / 1,200 (4.3%) | 0.2 |

<sup>1</sup>Median (Q1, Q3); n / N (%)
<sup>2</sup>Wilcoxon rank sum test; Pearson's Chi-squared test
<sup>3</sup>Percentages for malaria diagnostic tests were calculated among patients with an available result for the respective test. Patients without an available result were excluded from the denominator.

**eTable 6:** Factors associated with MTZ prescription within healthcare sectors in public and private healthcare facilities in South Benin, 2021: subgroup analysis.

| Variables | Public sector |  | Private sector |  |
| --- | --- | --- | --- | --- |
|  | aOR (95% CI) | p-value | aOR (95% CI) | p-value |
| Fever |  |  |  |  |
| No | — |  | — |  |
| Yes | 0.54 (0.34, 0.84) | 0.007 | 0.75 (0.52, 1.08) | 0.13 |
| Digestive symptoms |  |  |  |  |
| No | — |  | — |  |
| Yes | 9.00 (5.63, 14.8) | <0.001 | 8.25 (5.78, 12.0) | <0.001 |
| Respiratory tract symptoms |  |  |  |  |
| No | — |  | — |  |
| Yes | 0.42 (0.15, 0.98) | 0.062 | 0.43 (0.23, 0.76) | 0.005 |
| Skin lesions |  |  |  |  |
| No | — |  | — |  |
| Yes | 3.04 (1.55, 5.87) | 0.001 | 2.08 (1.22, 3.50) | 0.006 |
| Genitourinary symptoms |  |  |  |  |
| No | — |  | — |  |
| Yes | 9.36 (3.33, 28.5) | <0.001 | 3.75 (1.15, 12.1) | 0.025 |
| Malaria |  |  |  |  |
| No | — |  | — |  |

| Variables | aOR (95% CI) | p-value | aOR (95% CI) | p-value |
| --- | --- | --- | --- | --- |
| Yes | 0.11 (0.06, 0.18) | <0.001 | 0.27 (0.18, 0.39) | <0.001 |

Abbreviations: CI = Confidence Interval, OR = Odds Ratio

